# Does susceptibility to novel coronavirus (COVID-19) infection differ by age?: Insights from mathematical modelling

**DOI:** 10.1101/2020.06.08.20126003

**Authors:** Ryosuke Omori, Ryota Matsuyama, Yukihiko Nakata

## Abstract

Among Italy, Spain, and Japan, the age distributions of novel coronavirus (COVID-19) mortality show only small variation even though the number of deaths per country shows large variation. To understand the determinant for this situation, we constructed a mathematical model describing the transmission dynamics and natural history of COVID-19 and analyzed the dataset of fatal cases of COVID-19 in Italy, Spain, and Japan. We estimated the parameter which describes the age-dependency of susceptibility by fitting the model to reported data, taking into account the effect of change in contact patterns during the outbreak of COVID-19, and the fraction of symptomatic COVID-19 infections. Our modelling study revealed that if the mortality rate or the fraction of symptomatic infections among all COVID-19 cases does not depend on age, then unrealistically different age-dependencies of susceptibilities against COVID-19 infections between Italy, Japan, and Spain are required to explain the similar age distribution of mortality but different basic reproduction numbers (*R*_0_). Variation of susceptibility by age itself cannot explain the robust age distribution in mortality by COVID-19 in those three countries, however it does suggest that the age-dependencies of i) the mortality rate and ii) the fraction of symptomatic infections among all COVID-19 cases determine the age distribution of mortality by COVID-19.

## Introduction

Since its emergence, coronavirus disease 2019 (COVID-19) has resulted in a pandemic and has produced a huge number of cases worldwide (World Health Organization, 2020). As of May 29, 2020, the number of confirmed cases in Italy was 231,139, with 237,141 in Spain, and 16,683 in Japan (World Health Organization, 2020). Of those infected, it has been reported that elderly individuals account for a large portion of fatal cases inducing a large heterogeneity in the age distribution of mortality (Dowd et al., 2020; Onder et al., 2020; Wu et al., 2020).

The expected value of mortality (the number of deaths, hereafter referred to as mortality) is calculated as the product of the number of cases and the mortality rate among cases (hereafter referred to as morality rate). As the background mechanism of the heterogeneity of mortality by age, the association of two epidemiological factors with mortality can be considered: i) the age-dependency of susceptibility to infection, which is related to the heterogeneity in the number of cases, and ii) the age-dependency of severity, which is related to the heterogeneity in the mortality rate, e.g. the rate of becoming symptomatic, severe, or fatal case among infected individuals. For the first factor, a high susceptibility for infection will generate a larger number of infections and result in an increase in fatal cases. The possibility of heterogeneity in susceptibility by age was pointed out by the analysis of epidemiological data reported from Wuhan, China (Lee et al., 2020; Wu et al., 2020; Zhang et al., 2020) and from Iceland (Gudbjartsson, 2020). For the second factor, an increase in severity will result in a higher mortality rate and subsequently a rise in the number of fatal cases. This assumption is also reasonable because elder age as well as the existence of comorbidities, which are likely with aging, have been reported as risk factors for severe COVID-19 infections (Bonanad et al., 2020; Guan et al., 2020; Liu et al., 2020; Shi et al., 2020; Verity et al., 2020; Zhou et al., 2020). Although not yet shown in relation to severe acute respiratory syndrome corona virus 2 (SARS Cov-2), which is the causal agent of COVID-19, the presence of age-dependent enhancement of severity has been suggested in SARS coronavirus by the analysis of the innate immune responses in the BALB/c mouse model (Baas et al., 2008; Chen et al., 2010; Roberts et al., 2005).

Additionally, it has been suggested that antibody-dependent enhancement (ADE) can contribute to the formation of the observed age-dependency of severity, as suggested in SARS and Middle East respiratory syndrome (MERS) cases (Arabi et al., 2016; Drosten et al., 2014; Tay et al., 2020; Tetro, 2020; Wan et al., 2018; Yang et al., 2005).

Interestingly, the age distribution of mortality by COVID-19 (the distribution of the proportion of deaths per age group among all deaths), is similar between Italy, Japan, and Spain, even though the number of deaths are quite different among them (Ministry of Health, Labor and Welware, 2020; Epicentro, Istituto Superiore di Sanità, 2020; Centro de Coordinación de Alertas y Emergencias Sanitarias, 2020). The large difference in the number of deaths between the countries suggests a large difference in their basic reproduction numbers, *R*_0_s. An independency between age distribution of mortality by COVID-19 and *R*_0_ is suggested. From this independency of age distributions of mortality from *R*_0_, it can be expected that the contribution of heterogeneity in susceptibility by age to forming the age distribution of mortality is small. That is because, as we will show in this paper, though the age-dependency of severity will naturally produce a proportional effect on the distribution of mortality and result in the formation of robust distributions, when the age-dependency of susceptibility forms the age distribution of mortality, the age distribution of mortality highly depends on *R*_0_ and shows variability.

To understand the background of robust age distribution of mortality with varied *R*_0_, we constructed a mathematical model describing the transmission dynamics of COVID-19 and analyzed the impact of age-dependent susceptibility on the age distribution of mortality. The heterogeneity in social contacts by age may also contribute to the age distribution of mortality. Our model took into account the heterogeneity in social contacts by age and country, and the effect of behavioral change outside of the household during the outbreak. We also estimated and compared the age-dependent susceptibility in Japan, Italy, and Spain to argue the existence of heterogeneity in susceptibility among age groups.

## Results

Our result shows variation of susceptibility among age groups measured by the exponent parameter *φ* can explain the age distribution of mortality by COVID-19 (figure 2(a)). However, the age distribution of mortality formed by the age-dependency of susceptibility is influenced by the value of *R*_0_ (figure 2(b)), which cannot explain the similarity in age distributions of mortality among Italy, Japan, and Spain. On the other hand, if susceptibility is constant among age groups, the impact of *R*_0_ is quite small on the age distribution of mortality (figure 3).

**Figure 1:**
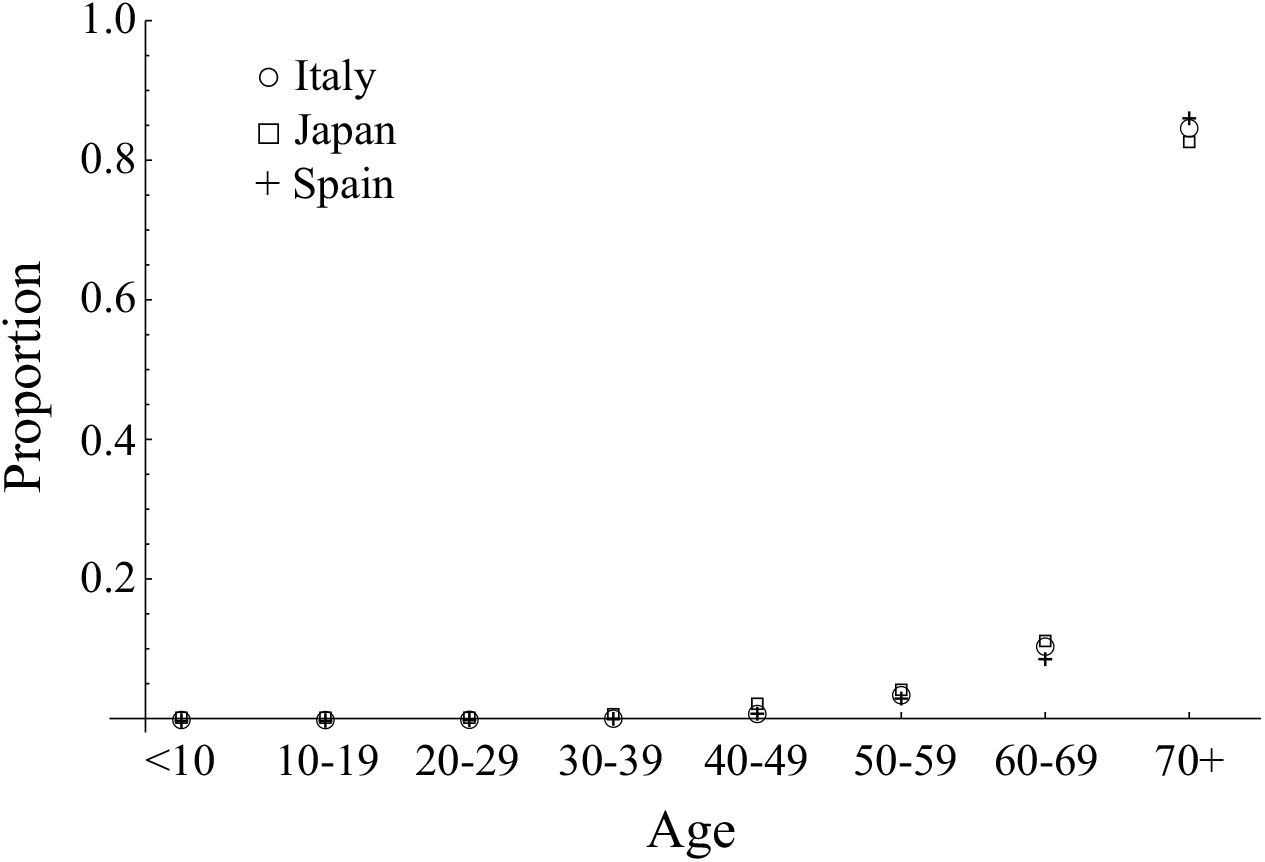
The age distribution of mortality by COVID-19 in Italy reported on 13th May 2020, Japan reported on 7th May 2020, and Spain reported on 12th May 2020. Circle, square, and “+” denote Italy, Japan, and Spain.

**Figure 2:**
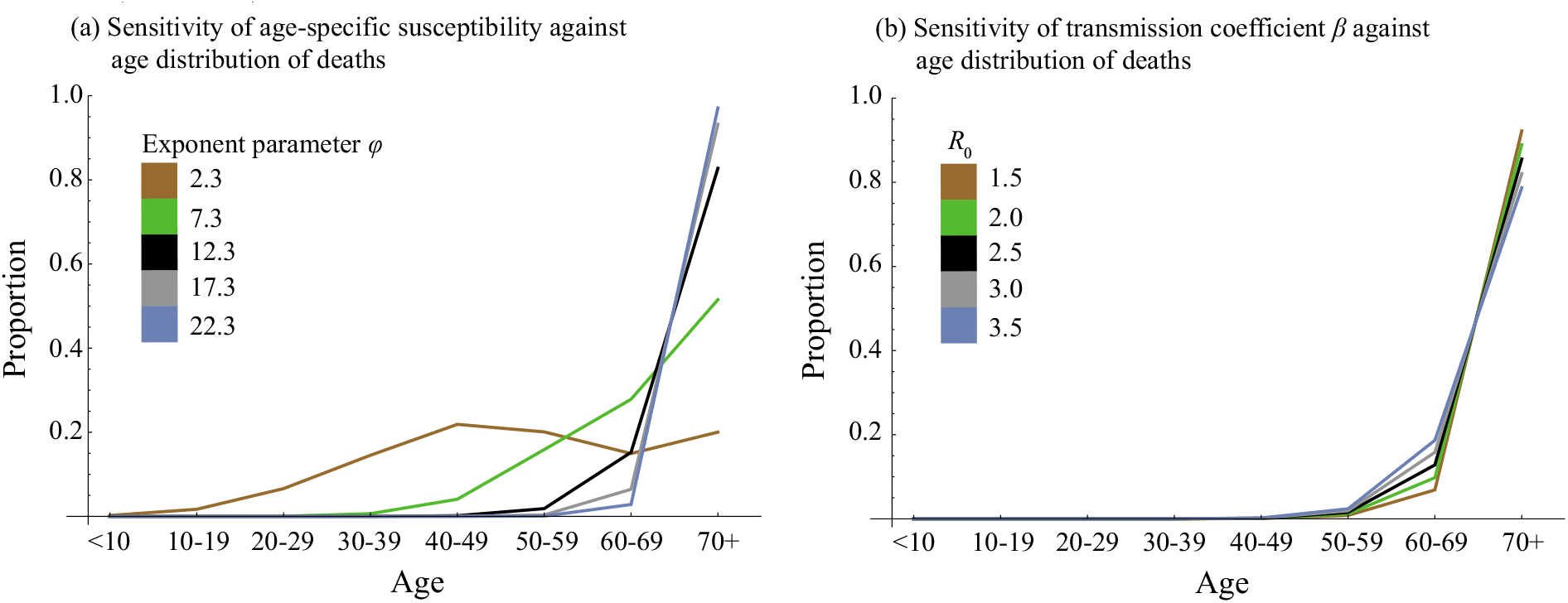
The sensitivity of (a) age-dependency of susceptibility and (b) transmission coefficient *β* against age distribution of mortality when age-independent mortality was assumed. In panel (a), all parameters except the exponent parameter *φ*, describing the variation of susceptibility among age groups, were fixed and parameterized as *R*_0_ = 2.9 in the setting for Spain. In panel (b), all parameters parameterized as the setting for Spain (*φ*=12.3) except were fixed except transmission coefficient *β*.

**Figure 3:**
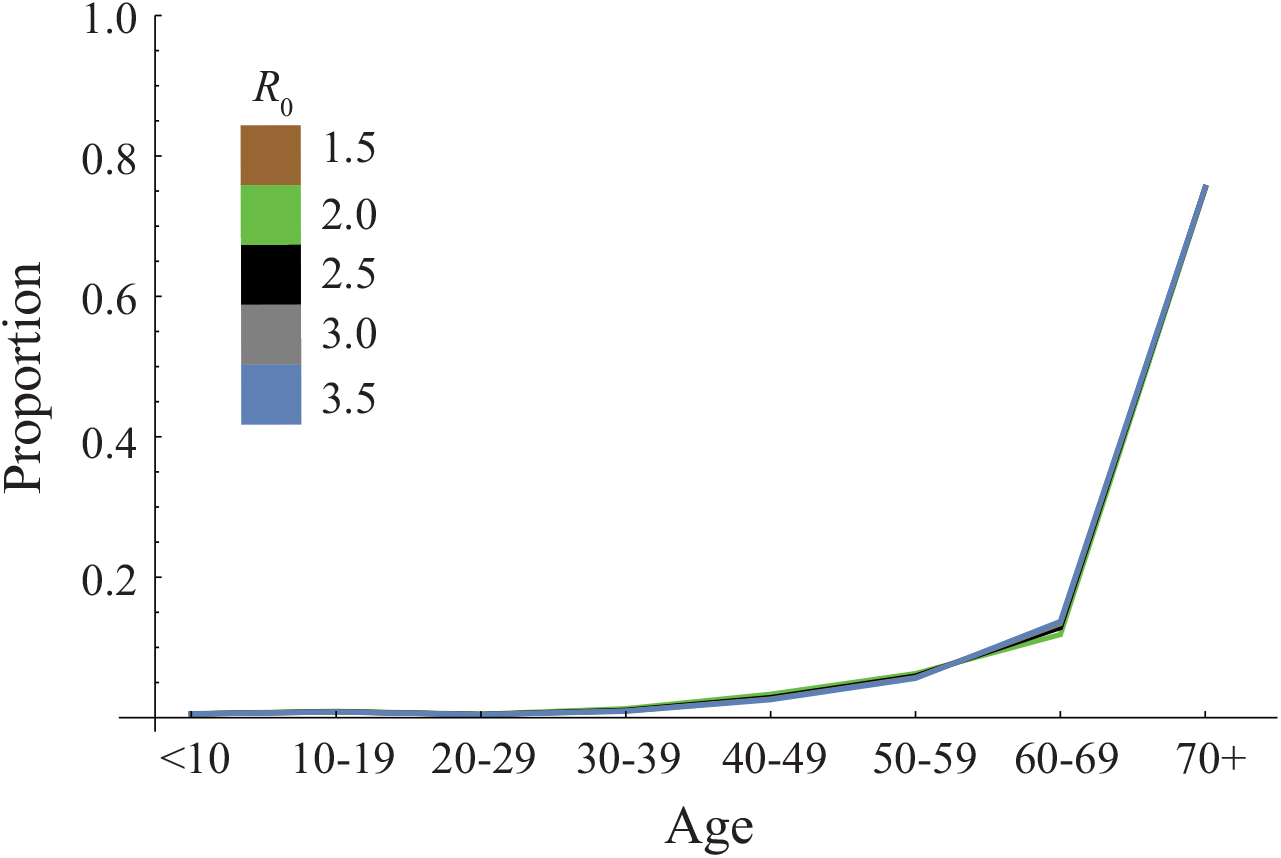
The sensitivity of transmission coefficient *β* against age distribution of mortality when it was assumed that age-dependent mortality was proportional to cCFR per age group. All parameters were fixed and parameterized as the setting for Spain except the transmission coefficient *β*.

Assuming that the age-dependency of mortality by COVID-19 is determined by only age-dependent susceptibility, i.e., the mortality rate does not depend on age, the exponent parameter, *φ*, describing the variation of susceptibility among age groups for each country, Italy, Japan, and Spain, was estimated as shown in figure 4. From the difference of the *R*_0_ value and country, the estimated value of *φ* is largely varied. The impact of reductions in contacts outside of the household on the estimated value of *φ* was small. The estimate of *φ* in Italy, assuming a range of *R*_0_ = 2.4-3.3 (Zhuang et al., 2020; D’Arienzo and Coniglio, 2020) was 15.0 (95% CI = 14.0-16.0), 16.3 (95% CI = 14.9-17.7), and 16.9 (95% CI = 15.4-18.4) for 80%, 40%, and no reduction in contacts outside of the household. For Japan, the estimate of *φ* assuming *R*_0_ = 1.7 (Expert Meeting on the Novel Coronavirus Disease Control, 2020) was 4.2 (95%CI = 3.7-4.9), 5.5 (95%CI = 4.9-6.3), and 6.1 (95%CI = 5.4-6.9) for 80%, 40%, and no reduction in contacts outside of the household. When it comes to Spain, the estimate of *φ* assuming an *R*_0_ = 2.9 (Caicedo-Ochoa et al., 2020) was 10.5 (95%CI = 10.4-10.6), 11.7 (95%CI = 11.6-11.9), and 12.3 (95%CI = 12.2-12.5) for 80%, 40%, and no reduction in contacts outside of the household.

**Figure 4:**
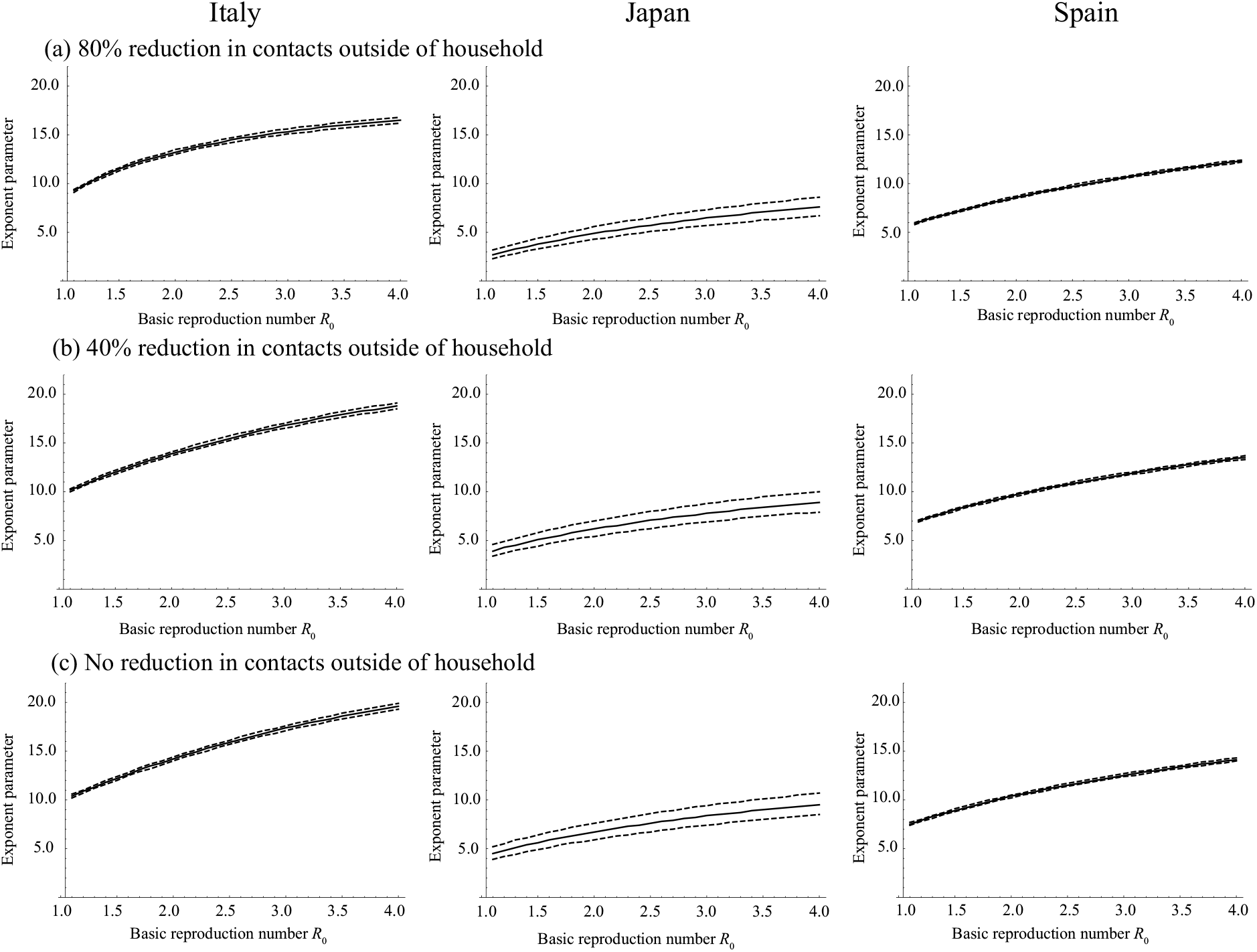
The estimate of exponent parameter *φ* describing the variation of susceptibility among age groups assuming that mortality rate does not depend on age. True and broken lines represent the maximum likelihood estimates and 95% confidence intervals, respectively.

The estimates of *φ*, assuming that the fraction of infections becoming symptomatic does not depend on age, were also varied by the value of *R*_0_ and by country (figure 5, 6 and 7). Employing the same assumptions of *R*_0_ value, the estimate of *φ* in Italy was 4.8 (95% CI = 4.2-5.3), 5.4 (95% CI = 4.9-5.9), and 5.7 (95% CI = 5.1-6.2) for 80%, 40%, and no reduction in contacts outside of the household. For Japan, the estimate of *φ* was 0.0 (95%CI = 0.0-0.9), 0.0 (95%CI = 0.0-1.1), and 0.0 (95%CI = 0.0-1.2) for 80%, 40%, and no reduction in contacts outside of the household. For Spain, the estimate of *φ* was 1.7 (95%CI = 1.4-1.9), 2.2 (95%CI = 1.9-2.5), and 2.5 (95%CI = 2.1-2.8) for 80%, 40%, and no reduction in contacts outside of the household.

**Figure 5:**
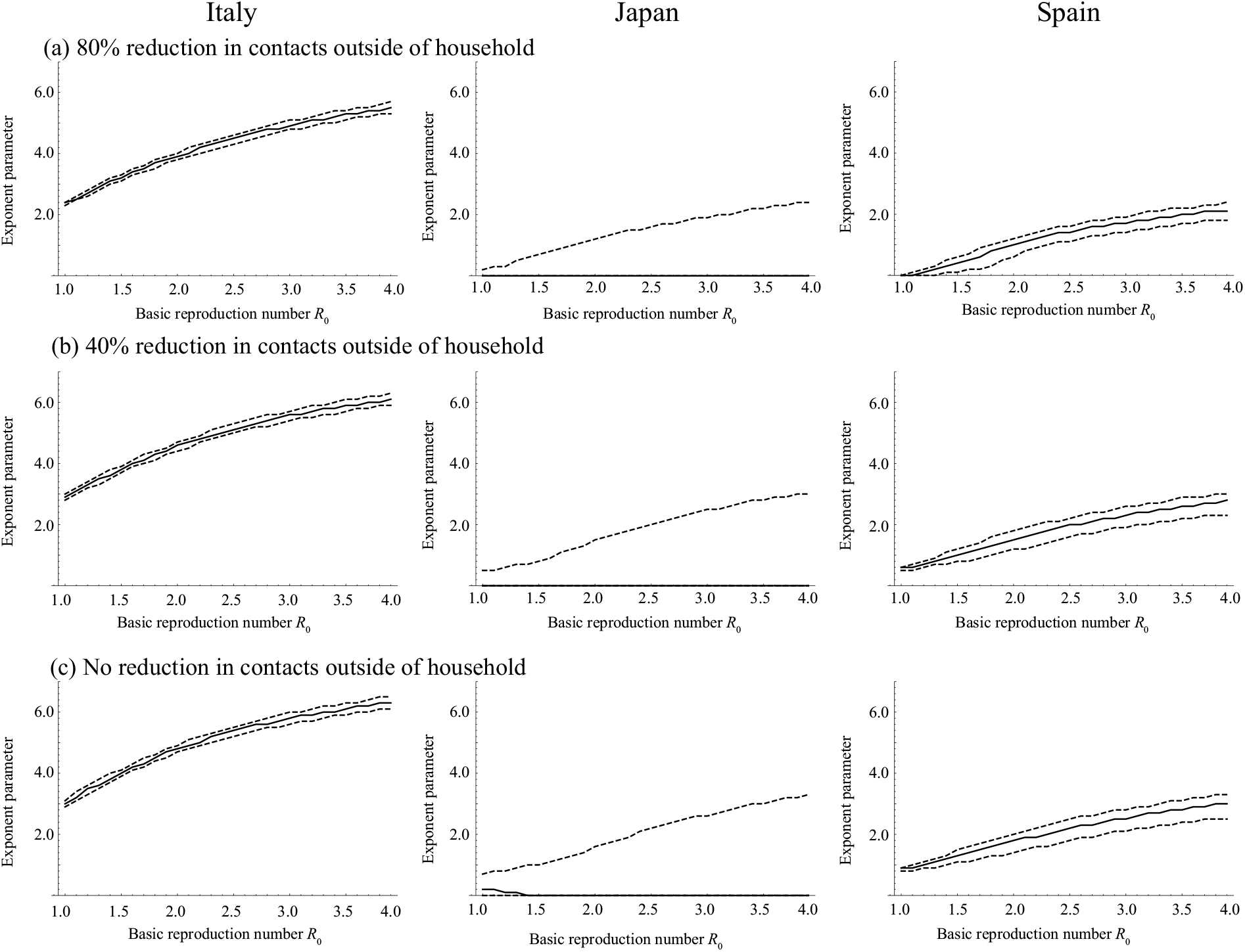
The estimate of exponent parameter *φ* describing the variation of susceptibility among age groups assuming that mortality rate does not depend on age and the fraction of infections that becomes symptomatic among all COVID-19 cases is 0.25. True and broken lines represent the maximum likelihood estimates and 95% confidence intervals, respectively.

**Figure 6:**
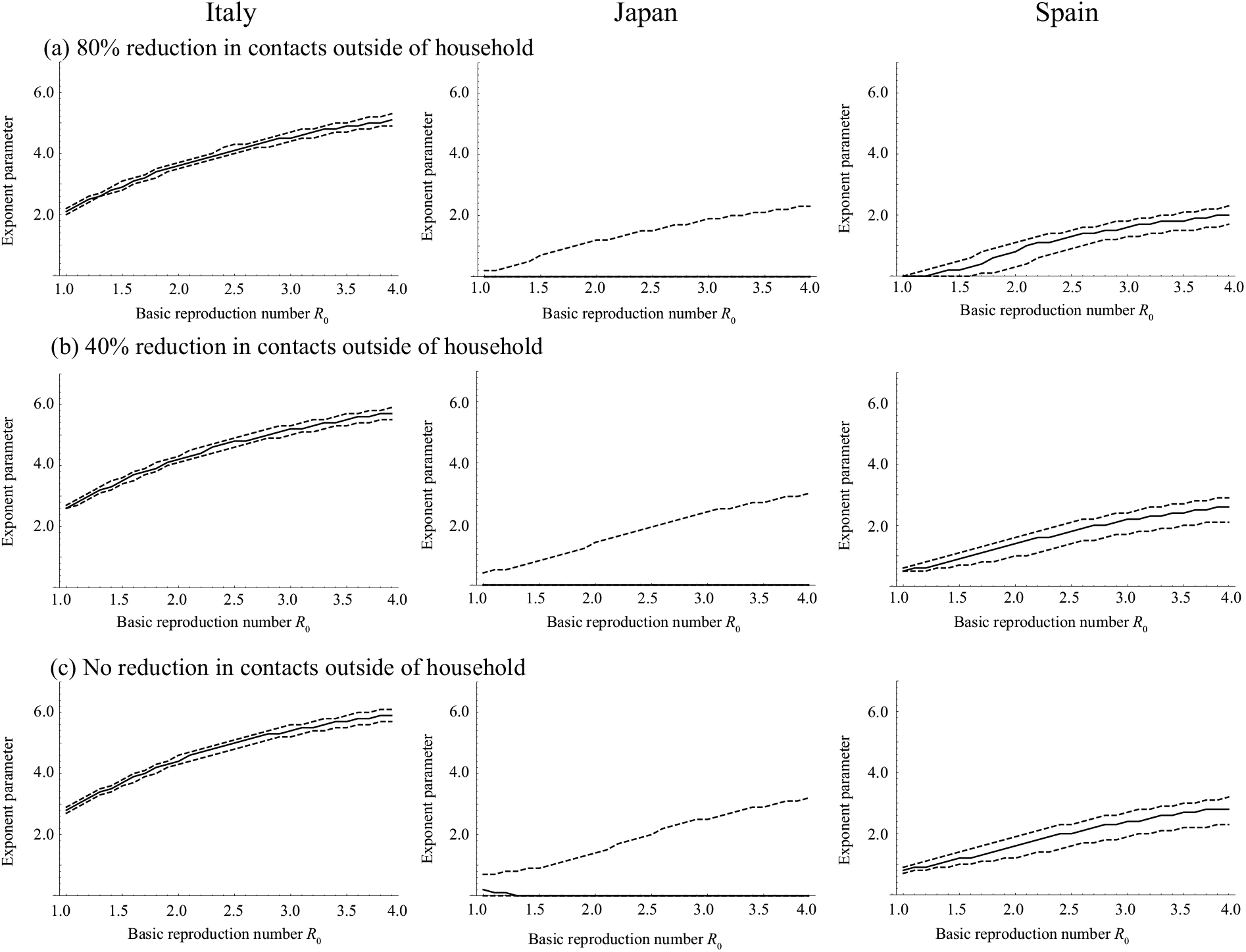
The estimate of exponent parameter *φ* describing the variation of susceptibility among age groups assuming that mortality rate does not depend on age and the fraction of infections that becomes symptomatic among all COVID-19 cases is 0.5. True and broken lines represent the maximum likelihood estimates and 95% confidence intervals, respectively.

**Figure 7:**
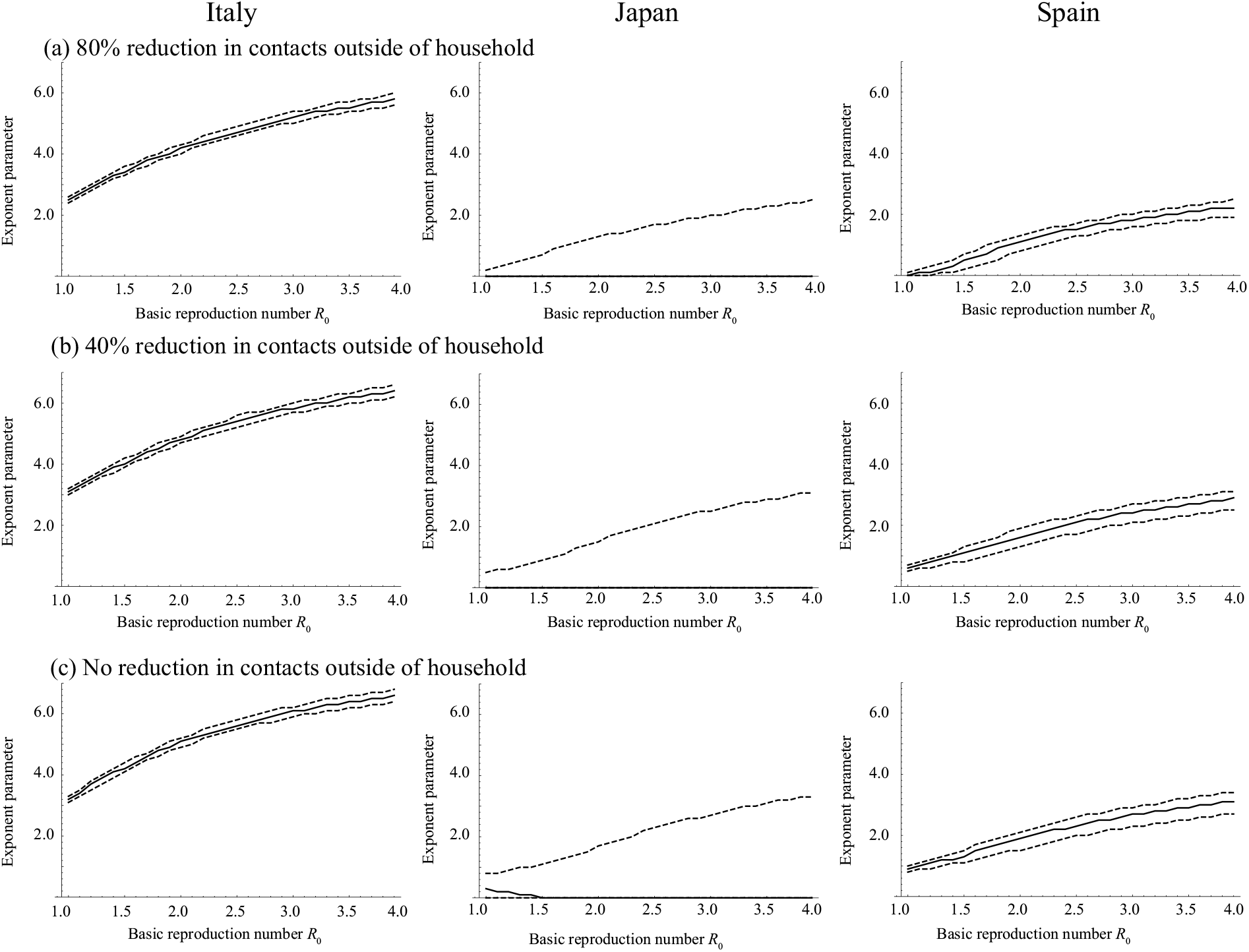
The estimate of exponent parameter *φ* describing the variation of susceptibility among age groups assuming that mortality rate does not depend on age and the fraction of infections that becomes symptomatic among all COVID-19 cases is 0.05. True and broken lines represent the maximum likelihood estimates and 95% confidence intervals, respectively.

## Discussion

In the present study, we explored the role of susceptibility to COVID-19 in explaining the age distribution of mortality by COVID-19. Interestingly, the age distributions of mortality from COVID-19 are quite similar between Italy, Japan, and Spain (figure 1). When comparing the age distributions of mortality, only the comparison between Italy and Spain is significant (*p*<0.05 in Wilcoxon rank sum test with Bonferroni correction). On the other hand, the numbers of deaths are quite different (29,525 for Italy, 400 for Japan, 18,818 for Spain). Indeed, *R*_0_ values are largely different: 2.4-3.3 for Italy (Zhuang et al., 2020; D’Arienzo and Coniglio, 2020), 1.7 for Japan (Expert Meeting on the Novel Coronavirus Disease Control, 2020), and 2.9 for Spain (Caicedo-Ochoa et al., 2020). If the variation of mortality by age is determined by only the age-dependency of susceptibility, the age distribution of mortality is affected by *R*_0_ as shown in figure 2(b). However, we observed a similarity in age distributions of mortalities between Italy, Japan, and Spain where their *R*_0_s are quite different. Indeed, unrealistically different *φ*s among these three countries are required to explain their age distribution of mortality for both settings, i) age-independent mortality, and, ii) the fraction of infections that becomes symptomatic among all COVID-19 cases, *f*_*s*_, does not depend on age. Although we cannot fully reject the existence of age-dependency in susceptibility, our results suggest that it does not largely depend on age, but rather that age-dependency in severity highly contributes to the formation of the observed age distribution in mortality.

The estimates of *φ*s assuming age independency in symptomatic infections were smaller than those that assumed age independency in mortality. This suggests that the age-dependency of the confirmed case fatality rate (cCFR), which can be biased by the age-dependent difference of the fraction of symptomatic infections among all cases, partially explains the age distribution in mortality. Indeed, when we assumed that the fraction of symptomatic infections was not dependent on age, the estimate of *φ* in Japan was close to zero in all scenarios regarding the fraction of symptomatic infections, meaning that susceptibility is constant among age groups (figure 5). Although we observed *φ*s around 5 in Italy and 2 in Spain, this does not mean straightforwardly that susceptibility is age dependent because there is room for an alternative explanation: not susceptibility, but an age-dependent fraction of symptomatic infections can explain this age-dependency. Unfortunately, as we do not yet have detailed data regarding the age-dependent fraction of symptomatic infections and the rate of diagnosis in COVID-19, we cannot conclude which factors (i.e., susceptibility or the fraction of symptomatic infection among all cases) contributed to the observed age-dependency.

Wu et al. (2020) showed variation of susceptibility to symptomatic infection by age. This susceptibility can be expressed as the product of the susceptibility and the fraction of symptomatic infection among all cases. To accurately understand susceptibility (i.e., without the constraint of the symptom onset), estimates of the age-dependent fraction of symptomatic infections is required.

To understand the mechanism of age-dependency of mortality by COVID-19, an accurate age-dependent mortality rate is required. To estimate the age-dependent mortality rate, an accurate estimate of the case fatality rate is required. However, the number of cases, which is the denominator of the case fatality rate, is difficult to estimate for COVID-19 due to changes in the testing rate (Gostic et al., 2020a; Gostic et al., 2020b; Omori et al. 2020), the change of case definition (Tsang et al. 2020), selection biases (Bar-on et al., 2020), and the delay between the onset of symptoms and death (Linton et al., 2020; Shim et al., 2020; Sun et al., 2020; Verity et al., 2020) as were the cases we experienced in the surveillance of other emerging diseases (Ghani et al., 2005; Garske et al,, 2008). To address this problem, implementation of active epidemiological surveillances, such as a large-scale cohort study including real-time detection of infections, should be considered.

From the modelling perspective on mortality by Covid-19, age-dependency of severity should be carefully taken into consideration. In particular, in the mathematical models of ADE, the previous models employed three types of assumptions (Woodall and Adams, 2014), the assumption of: increasing susceptibility to infection (Recker et al., 2009; Tang et al., 2018), increasing transmissibility once infection occurred (Ferguson et al., 1999; Ferguson and Andreasen, 2002; Recker et al., 2009), and increasing severity and/or mortality associated with infection (Kawaguchi et al., 2003). Based on our results and from the biological/epidemiological observations of past SARS and MERS cases, the “increasing severity” assumption should be taken into account when analyzing SARS Cov-2 epidemics.

In conclusion, the contribution of age-dependency to susceptibility is difficult to use to explain the robust age distribution in mortalities by COVID-19, and it suggests that the age-dependencies of the mortality rate and the fraction of symptomatic infections among all COVID-19 cases determine the age distribution in mortality from COVID-19. Further investigations regarding age-dependency on the fraction of infections becoming symptomatic is required to understand the mechanism behind the mortality by COVID-19 infections.

## Methods

### 1. Data

We analyzed the number of mortalities caused by COVID-19 in Italy reported on 13th May 2020, Japan reported on 7th May 2020, and Spain reported on 12th May 2020. The data were collected from public data sources in each country (Ministry of Health, Labor and Welware, 2020; Epicentro, Istituto Superiore di Sanità, 2020; Centro de Coordinación de Alertas y Emergencias Sanitarias, 2020).

### 2. Model

To understand the background of robust age distribution of mortality with varied *R*_0_, we employed a mathematical model describing transmissions of COVID-19, an age-structured SEIR model, which can be written as;

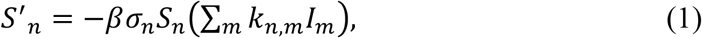

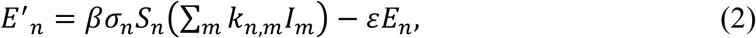

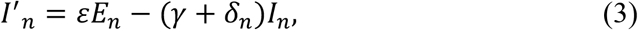

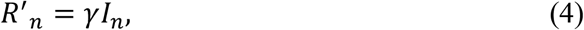

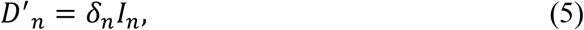

where *S*_*n*_, *E*_*n*_, *I*_*n*_, *R*_*n*_ and *D*_*n*_ represent the proportion of susceptible, latent, infectious, recovered and dead among the entire population, and the subscript index *n* denotes age group. We stratified the entire population by into eight groups, *n* = 1, 2, 3, 4, 5, 6, 7, and 8 for < 10 years old (yo), 10-19 yo, 20-29 yo, 30-39 yo, 40-49 yo, 50-59 yo, 60-69 yo, and 70+ yo. *β, k*_n,m_, *ε, γ* and *δ*_*n*_ represent a transmission coefficient, an element of the contact matrix between age group *n* and *m*, the progression rate from latent to infectious, recovery rate and mortality rate among age group *n*, respectively. *σ*_n_ denotes the susceptibility of age group *n*. For the sake of simplicity, births and deaths by other than COVID-19 were ignored. To take into account the effect of behavioral changes outside of the household during the outbreak, *k*_n,m_ is decomposed by a matrix for contacts within household *k*_in,n,m_ and that for contacts outside the household *k*_out,n,m_;

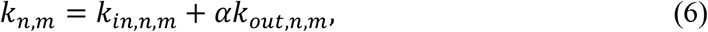

where *α* denotes the reduced fraction of contacts outside of the household. We modelled age specific susceptibility as

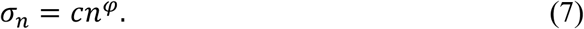

Where *c* is a constant among all age groups, *φ* denotes the exponent parameter describing the variation of susceptibility among age groups. An increase in *φ* means an increase in the variation of susceptibility among age groups, and *φ*=0 means that susceptibility is equal among all age groups

### 3. Parameterizations

We parameterized *ε* and *γ* using the values from a previous modelling study of COVID-19 (Prem et al., 2020). We referred to the contact matrices for Italy, Japan, and Spain from Prem et al. (Prem et al., 2017). *β* and *c* were controlled such that the basic reproduction number, *R*_0_, becomes arbitral values. *R*_0_ was calculated by constructing a next generation matrix (Diekman and Heesterbeek, 2000; Mossong et al., 2008) using each country’s demographic data obtained from a public data source (United Nations, 2020).

### 4. Fitting

We calculated the proportions of deaths in the age group *n* among all deaths, *d*_n_ (= *D*_*n*_(∞)/ ∑_*n*_ *D*_*n*_(∞)), and fitted them to the observed data in each country. The mortality rate among age group *n, δ*_*n*_, is required to calculate *d*_n_, however, a reliable estimate of *δ*_*n*_ for COVID-19 is difficult to obtain. Due to the uncertainty of the fraction of symptomatic infections per age group, *δ*_*n*_ is difficult to estimate from observed data, i.e., the confirmed case fatality rate among age group *n* (cCFR_*n*_). Since an estimate of *δ*_*n*_ is difficult to obtain, we employed two different settings to calculate *d*_n_, i) *δ*_*n*_ is assumed to be a constant among all age groups, or, ii) *δ*_*n*_ is calculated from cCFR_*n*_ assuming that the fraction of symptomatic infections among all COVID-19 cases (*f*_*s*_) is not dependent with age.

In setting i), the value of *δ*_*n*_ is not required to estimate *d*_n_ once the value of *R*_0_ is given. We calculated *d*_n_ by calculating the proportions of recovered persons per age group among all recovered persons *R*_*n*_(∞)/ ∑_*n*_ *R*_*n*_(∞) instead of *D*_*n*_(∞)/ ∑_*n*_ *D*_*n*_(∞). In our model, shown in equation (1)-(4), *R*_*n*_(∞)/ ∑_*n*_ *R*_*n*_(∞) is determined by the value of *R*_0_ completely when all parameter values other than *β* and *δ*_*n*_ are fixed, and *D*_*n*_(∞)/ ∑_*n*_ *D*_*n*_(∞) = *R*_*n*_(∞)/ ∑_*n*_ *R*_*n*_(∞) if *δ*_*n*_ ≠0. The proof can be found in supplementary file 1.

The assumption in setting i), *δ*_*n*_ is constant among all age groups, may be too strong for the COVID-19 epidemic. To take into account the age-dependency of mortality by COVID-19, *δ*_*n*_ was calculated from the cCFR_*n*_ assuming that *f*_*s*_ is not dependent with age. As for the setting ii), assuming three scenarios; *f*_*s*_ = 0.05, 0.25, and 0.5, *δ*_*n*_ for each country were calculated using cCFR_*n*_ in each country. We obtained *δ*_*n*_ by solving cCFR_*n*_ = *δ*_*n*_/[*f*_*s*_ (*δ*_*n*_+*γ*)].

We solved the model shown in equations (1)-(5) numerically, and *d*_n_ was calculated after sufficient time was given to finish the epidemics. We estimated *φ* using a log likelihood function describing the multinomial sampling process of deaths per age group;

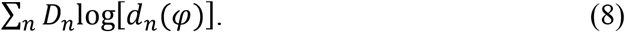

Maximum likelihood estimates of *φ* with given *R*_0_ were obtained by maximizing equation (8) and the profile likelihood-based confidence intervals were computed.

## Data Availability

All relevant data are within the manuscript and Supporting Information files.

## Acknowledgement

The authors gratefully acknowledge the manuscript language review of Dr Heidi L. Gurung.

## Ethical approval

This study was based on publicly available data and did not require ethical approval.

